# Clinical Spectrum, Treatments and Outcomes of VEXAS Syndrome: A Multicenter Belgian Cohort

**DOI:** 10.64898/2026.08.26.26361409

**Authors:** L. Funaro, L. Naesens, A. Betrains, B. Vokaer, B. Couturier, O. Malaise, G. Vertenoeil, F. Lambert, R. Lattenist, F. Vandergheynst, L. Wolff

## Abstract

**Background:** VEXAS syndrome is a late onset autoinflammatory disease caused by somatic *UBA1* mutations and characterized by heterogeneous systemic and hematologic manifestations. We aimed to describe all identified Belgian cases through a national multicenter cohort.

**Methods:** We conducted a retrospective study across four Belgian tertiary centers. Clinical, biological, genetic, therapeutic, and outcome data were collected using standardized anonymized case report forms. Analyses were descriptive.

**Results:** Twenty-one male patients were identified between January 2018 and May 2025. General symptoms such as Fatigue, weight loss and sweating occurred in 95% of cases. The most frequent manifestations were cutaneous (85.7%), hematologic (76.2%), articular (66.7%), thromboembolic (57.1%), chondritis (42.9%), ophthalmologic (38.1%), pulmonary (38.1%). Other manifestations also included vasculitis (61.9%). At diagnosis, 95% had anemia, macrocytic in 57%, and 28.6% had thrombocytopenia. Corticosteroids were the main first-line therapy. Second-line treatments included anti–IL-6 agents (46.7%), JAK inhibitors (20%), and azacitidine (14.3%). Complete remission occurred in 50% of patients receiving anti–IL-6 therapy and in 33% treated with either JAK inhibitors or azacitidine. Two patients underwent allogeneic stem cell transplantation, one died from infectious complications. Twenty-six infectious episodes were recorded, including opportunistic infections. Six patients (28.6%) died during follow-up, four from infectious complications.

**Conclusion:** This first Belgian national cohort confirms the clinical heterogeneity of VEXAS syndrome and highlights substantial infectious morbidity and mortality. Access to targeted second-line therapies, particularly anti–IL-6 agents and JAK inhibitors, remains challenging despite apparent clinical benefit.

## Introduction

In 2020, VEXAS (Vacuoles, E1 enzyme, X-linked, auto-inflammatory and somatic) discovery marked a turning point for auto-inflammatory disease caused by a somatic mutation in *UBA1* gene. The identification of this syndrome challenged the traditional paradigm that autoinflammatory diseases are exclusively caused by germline mutations (1). This condition manifests with old males presenting with various systemic and immunological features, including general symptoms such as fever, asthenia, weight loss, as well as cutaneous, articular, pulmonary, and thromboembolic complications often associated with hematological abnormalities, most commonly myelodysplastic syndromes (2)(3). Treatment mainly focuses on symptom control. First-line therapy consists of corticosteroids, but patients are usually steroid-dependent and require high doses. This justifies the use of second line treatments such as JAK inhibitors and anti–IL-6R antibodies, or hypomethylating agents targeting the pathological clone, such as azacitidine (5). The only curative option relies on allogeneic stem cell transplantation (6). As VEXAS is a newly described entity with a poor prognosis that often requires complex treatments whose accessibility varies from one country to another, it is important to describe national experiences. Considering the uncertainty surrounding prophylactic strategies and the high infectious morbidity and mortality, a focus was given to microbiological data (7). We describe in this article clinical manifestations, biological data, treatments and outcomes in patient diagnosed with VEXAS syndrome across Belgium.

## Methods

We conducted a retrospective study across four Belgian centers: the Hôpitaux Universitaires de Bruxelles (H.U.B.) Erasme Hospital and Jules Bordet Institute, the University Hospital of Ghent, the Hôpital Universitaire de Liège, and the University Hospital of Leuven. All patients with a genetically confirmed *UBA1* mutation were included, regardless of the sequencing method used. This study was approved by the Ethics Committee of HUB (reference: P2025/015) as well as by the respective committees at each participating center. A REDCap tool was implemented in order to create a centralized database. Data were anonymized and collected using a Case Report Form (CRF), which was completed in each center by the referring practitioner. The CRF captured information on various clinical manifestations across the different systemic systems, as well as relevant biological parameters. Special attention was given to treatments, treatment response, and infectious events. This study was conducted in accordance with the Strengthening the Reporting of Observational Studies in Epidemiology (STROBE) guidelines. We performed a descriptive analysis of our sample. Qualitative variables were expressed as percentages. The normality of distributions was assessed using the Shapiro-Wilk test. Variables following a normal distribution were described using means and standard deviations, while non-normally distributed variables were described using medians and interquartile ranges. Complete remission was defined by the absence of symptoms attributed to VEXAS syndrome, a CRP (C-reactive protein) lower than 10 mg/L, and a corticosteroid dose of less than the equivalent of 10 mg of prednisone per day (8). Biological data were collected from the closest date of the diagnosis corresponding at the genetic analysis showing evidence of the *UBA1* mutation.

## Results

Thirty-four patients were screened carrying a *UBA1* mutation but, due to a lack of data, only 21 patients were included in the study. All were male, and the majority (18/21) were Caucasian. The mean age at first manifestation was 67.8 ± 9.5 years. The median diagnostic delay was 14 months (IQR 16). Constitutional symptoms were frequent: fever of unknown origin occurred in 10/21 patients (47.6%), asthenia in 20/21 (95.2%), anorexia in 13/21 (61.9%), and night sweats in 9/21 (42.9%). Weight loss was reported in 18/21 patients (85.7%), with a mean loss of 7.0 ± 3.8 kg (approximately 2.6 kg/month). All patients had at least one organ system involved, predominantly cutaneous (85.7%), hematologic (76.2%), articular (66.7%), and thromboembolic events (57.1%) (Table 1). Cutaneous manifestations included neutrophilic dermatosis in 5/21 patients (23.8%), leukocytoclastic vasculitis in 5/21 (23.8%), erythema nodosum in 3/21 (14.3%), and urticarial vasculitis in 2/21 (9.5%). Three patients (14.3%) presented non-specific eruptions on histopathology sections. Hematologic involvement occurred in 16/21 patients (76.2%). myelodysplastic syndrome (MDS) was present in 11/21 (52.4%). MDS subtypes included multilineage dysplasia (3/21, 14.3%), single-lineage dysplasia (1/21, 4.8%), and MDS not otherwise specified with ring sideroblasts (1/21, 4.8%). No high-risk MDS was observed at diagnosis, although one patient progressed to MDS with excess blasts (IPSS intermediate-1) after one year. Monoclonal gammopathy of undetermined significance (MGUS) was identified in 3/21 patients (14.3%), and one patient (4.8%) had lymphoplasmacytic lymphoma. Seventeen bone marrow examinations were available. Vacuolization was present in 12/17 (70.5%) and dysplasia in 12/17 (70.5%) on bone marrow smears, while bone marrow fibrosis was identified in 2/17 (11.8%) patients on biopsy. No excess of blasts was detected at baseline. Articular manifestations occurred in 14/21 patients (66.7%). Arthralgia was universal, and 6/21 (28.6%) had polyarticular involvement (>5 joints). Ankles (28.6%) and hands (19.1%) were most frequently affected. Arthritis was documented in 4/21 patients (19,0%). Chondritis occurred in 9/21 cases (42.9%), mainly involving the ears (38%) and nose (14%). Thromboembolic events were observed in 12/21 patients (57.1%), most commonly deep venous thrombosis (33.3%) and superficial venous thrombosis (28.6%). Vasculitis occurred in 13/21 patients (61.9%): large-vessel involvement in 5/21 (23.8%) and medium-vessel involvement in 3/21 (14.3%) and Small vessel involvement in 8/21 (38,1%). Respiratory involvement was present in 8/21 patients (38.1%), most frequently ground-glass opacities (3/21, 14.3%) on CT-scan imaging. Pulmonary infiltrates and consolidations were also observed in 2/21 patients (9.5%), and one patient had pleural effusion. Ophthalmologic manifestations occurred in 8/21 (38.1%), including episcleritis in 5/21 (23.8%) and anterior uveitis in one case. Lymphadenopathy was detected in 8/21 patients (38.1%), mainly cervical/axillary (19%), iliac/mediastinal (14.3%), and inguinal (9.5%).

**Table 1.**
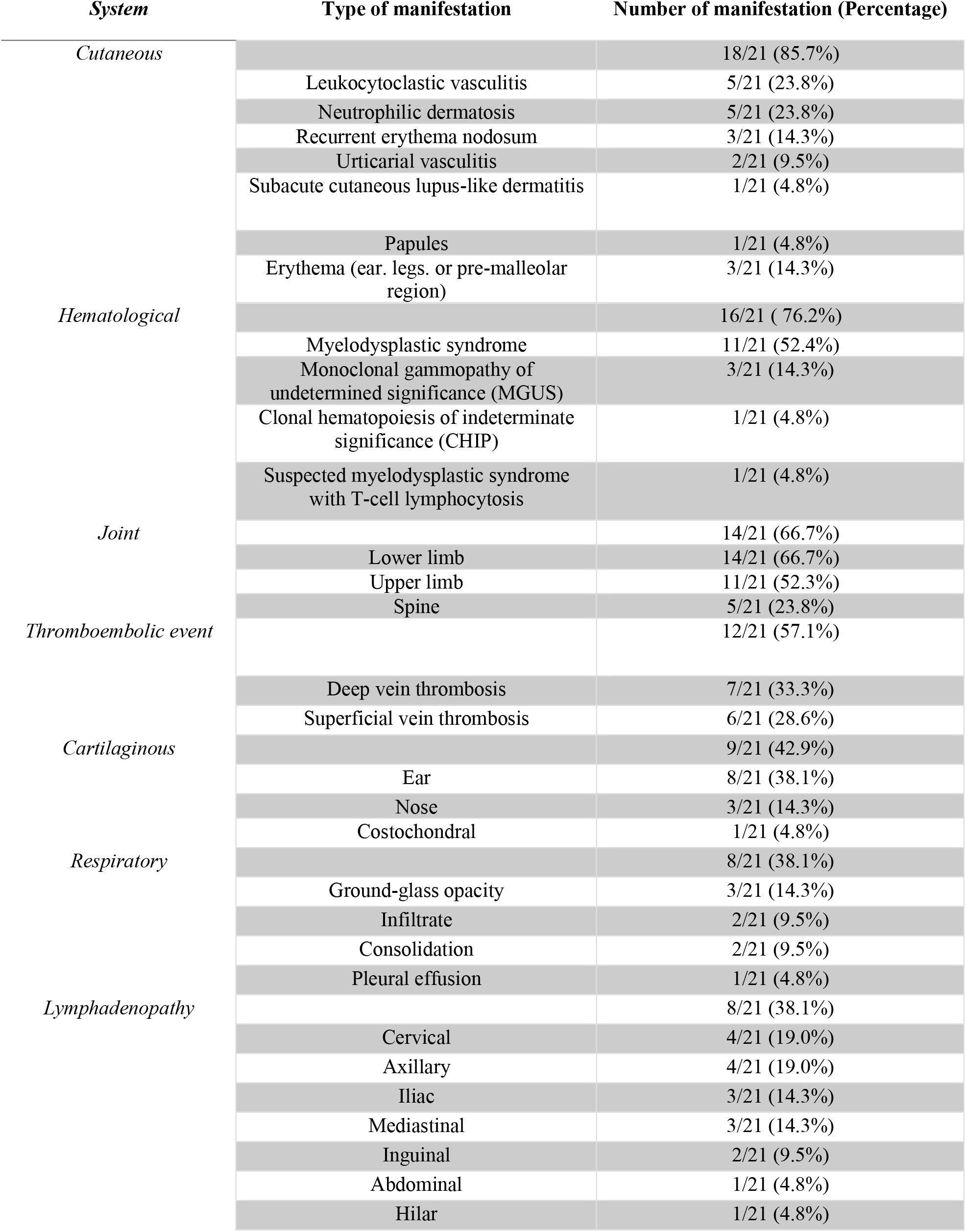
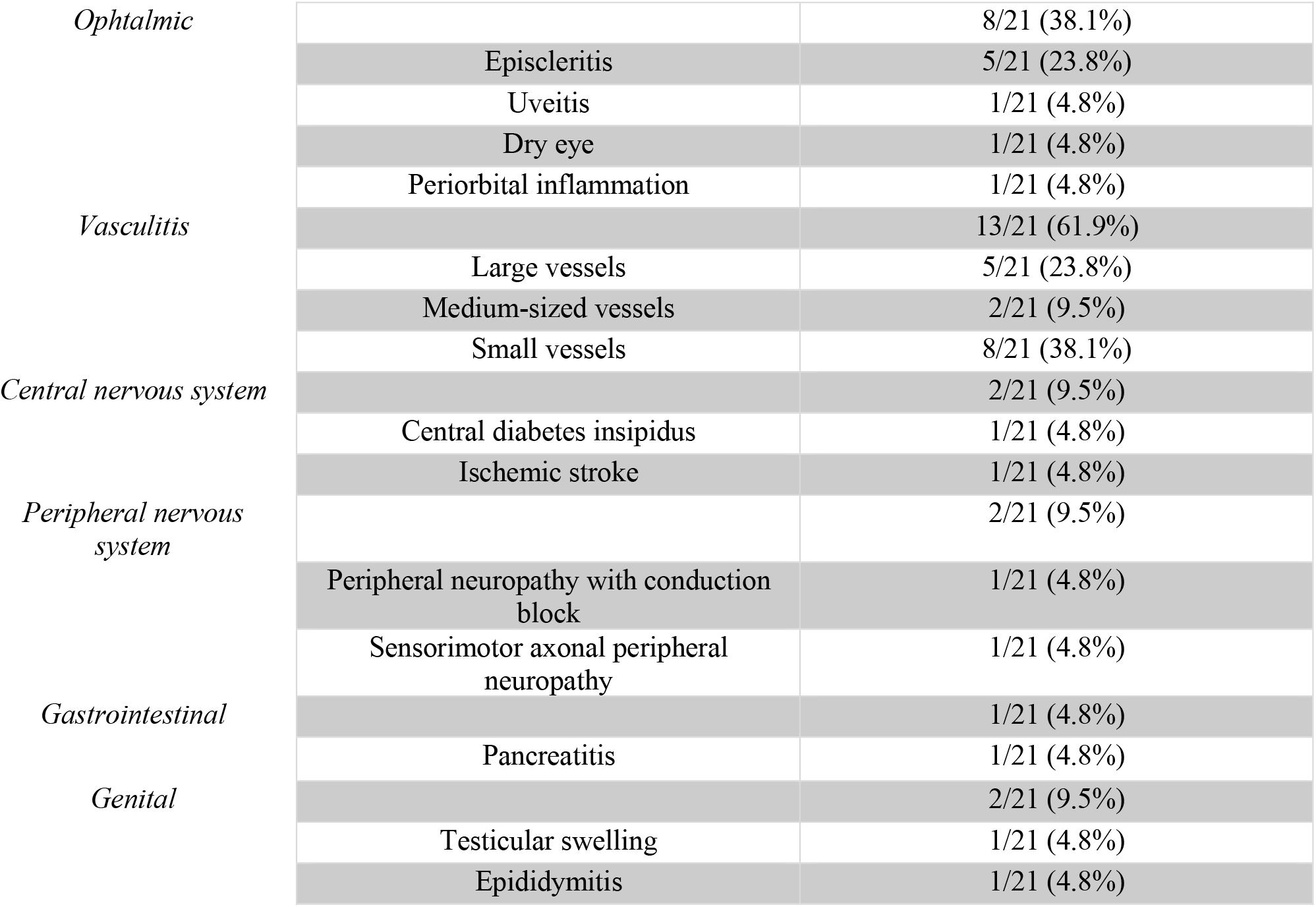
Manifestations per systems in belgian VEXAS patients.

Regarding genotype, 14 patients (66.7%) carried the p.(Met41Thr) mutation, three (14.2%) p.(Met41Leu), two (9.5%) p.(Met41Val), and one (4.8%) p.(Ser51Phe). One patient carried p.(Ser56Phe). Mean variant allele fraction was 58.4 ± 20.8%. The p.(Ser56Phe) patient uniquely had normal CRP at diagnosis and presented with MDS and marrow fibrosis. Among the cohort, the patient with the p.(Ser56Phe) variant was the only one with normal CRP at diagnosis and presented with MDS associated with marrow fibrosis. At diagnosis, anemia was present in 20/21 patients (95.2%) with a mean hemoglobin of 10.1 ± 1.8 g/dL (Table 2). It was macrocytic in 12/21 (57.1%), normocytic in 7/21 (33.3%), and microcytic in one patient with concomitant thalassemia. Mean MCV was 102 ± 10 fL. Thrombocytopenia occurred in 6/21 patients (28.6%) and thrombocytosis in one. Mean platelet count was 208 ± 106 G/L. Median leukocyte count was 4.8 G/L (2.8–6.8) with one case of mild neutropenia. Lymphopenia occurred in 12 patients. CRP was elevated in 20/21 (95%) with a mean value of 94.3 ± 53 mg/L. ANA were detected in 3/21 patients (≥1:160), speckled in two cases. ANCA positivity was observed in five patients without PR3 or MPO specificity.

**Table 2.** Median, mean and proportions of biological parameters at diagnosis from Belgian VEXAS patients.

| *Parameter* | *Values* |  |
| --- | --- | --- |
| *Hemoglobin (g/dL)* | 10.1 (±1.8) |  |
| *Mean corpuscular volume (MCV, fL)* | 102 (±10.0) |  |
| *Platelets (×10<sup>9</sup>/L)* | 208 (±106) |  |
| *White blood cells (WBC, ×10<sup>9</sup>/L)* | 4.8 (2.8-6.8) |  |
| *Neutrophils (×10<sup>9</sup>/L)* | 2.8 (1.1-4.5) |  |
| *Lymphocytes (×10<sup>9</sup>/L)* | 1.27 (±0.6) |  |
| *Eosinophils (×10<sup>9</sup>/L)* | 0.05 (0.02-0.1) |  |
| *C-reactive protein (CRP, mg/L)* | 94.3 (±53) |  |
| *Serum creatinine (mg/dL)* | 0.8 (0.6-1.1) |  |
| *Prothrombin time (PT, s)* | 47 (±31.3) |  |
| *International normalized ratio (INR)* | 1.2 (±0.1) |  |
| *Activated partial thromboplastin time (aPTT, s)* | 30.6 (±6.4) |  |
| *Fibrinogen (g/L)* | 6.45 (4.3-8.7) |  |
| *Anemia* |  | 20/21 (95.2%) |
|  | Macrocytic | 12/21 (57.1%) |
|  | Normocytic | 7/21 (33.3%) |
|  | Microcytic | 1/21 (4.7%) |
| *Thrombocytopenia* |  | 6/21 (28.6%) |
| *Lymphopenia* |  | 8/21 (38.1%) |

First-line therapy consisted of corticosteroids in 20/21 patients (95.2%); one patient received clone-targeting therapy (lenalidomide and azacitidine). Seventeen patients (81%) required second-line treatment, most frequently anti-IL-6 agents (23.8%) or JAK inhibitors (9.6%) (Table 3). Patients received a mean of 3 ± 1.5 treatment lines, with a median time to next treatment of 4 months (0.5– 7.5). At study end, 10/21 patients (47.6%) were symptom-free, 9/21 (42.9%) remained corticosteroid-dependent, and 5/21 (23.8%) achieved complete remission (Table 4).

**Table 3.**
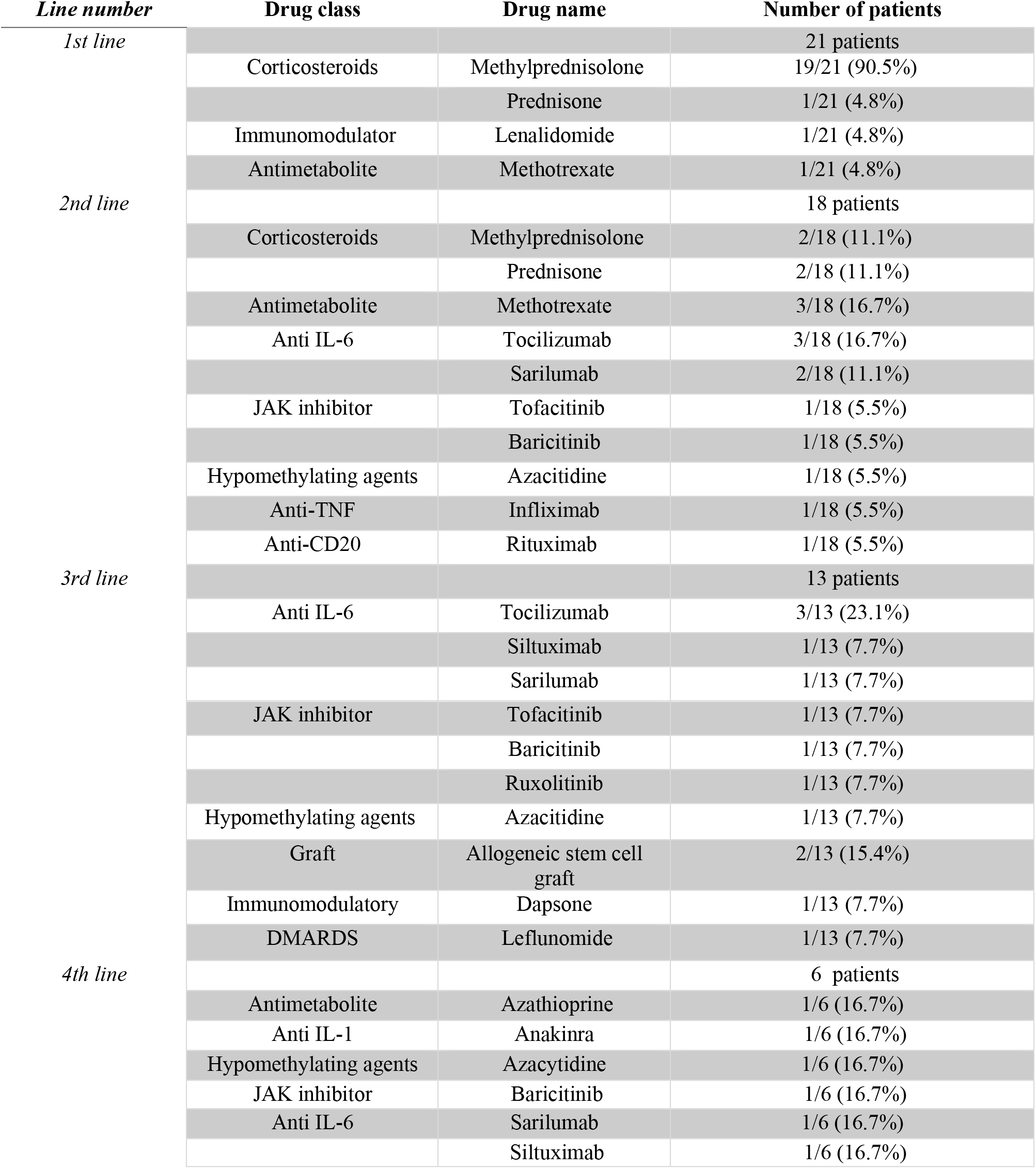
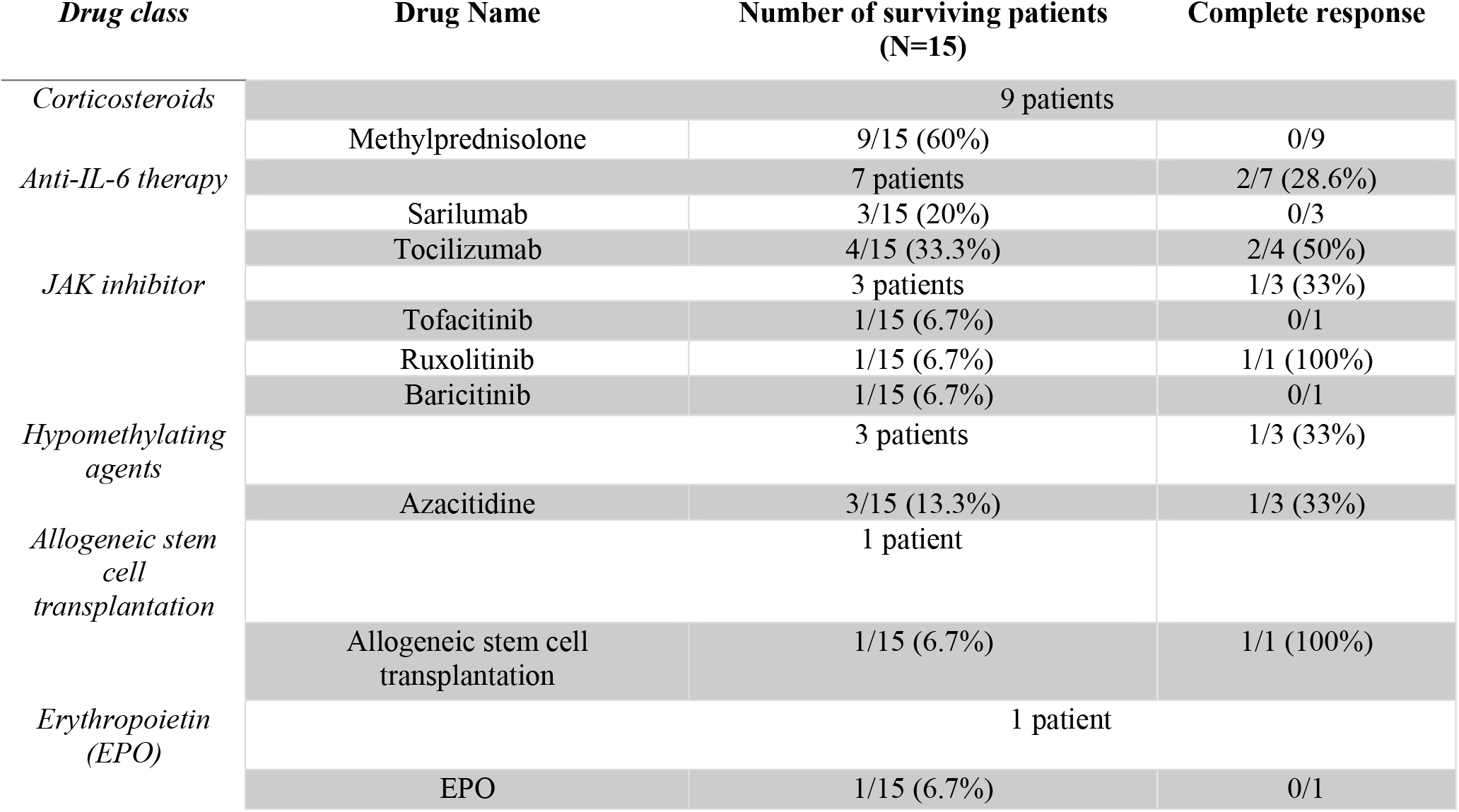
Different lines of treatment during follow-up (IL-6: Interleukin-6; JAK: Janus kinase; TNF: Tumor necrosis factor; CD20: Cluster of Differentiation 20; IL-1: Interleukin-1, DMARD : disease-modifying antirheumatic drug).

**Table 4.** Treatment resulting in complete remission among our surviving patients at the end of the study. (IL-6: Interleukin-6 receptor; JAK: Janus kinase). Complete remission was defined by the absence of symptoms attributed to VEXAS syndrome, a CRP (C-reactive protein) lower than 10 mg/L, and a corticosteroid dose of less than the equivalent of 10 mg of prednisone per day.

Twelve patients experienced at least one documented infection. Among 26 infectious episodes, 17 (65.4%) were bacterial, 8 (30.8%) viral, and 3 (11.5%) fungal counting surinfection with other pathogens as a single episode (Table 5). Common pathogens included *Escherichia coli, Pseudomonas aeruginosa*, SARS-CoV-2, and *Aspergillus fumigatus*. One patient received prophylaxis with trimethoprim-sulfamethoxazole. The lungs were the most frequently affected site (46%), followed by bacteremia (26.9%), and renal or cutaneous infections (7.7%each) (Table 6). Sixteen infections were complicated by respiratory failure or necrotizing pneumonia. Sepsis occurred in 5/26 (19.2%) and septic shock in 4/26 (15.3%). Four infection-related deaths were recorded, including one invasive aspergillosis. Overall, 19 infections resolved (73.1%). At study completion, 6/21 patients (29%) had died: four from infection, one from ST-elevation myocardial infarction, and one from an undetermined cause.

**Table 5.**
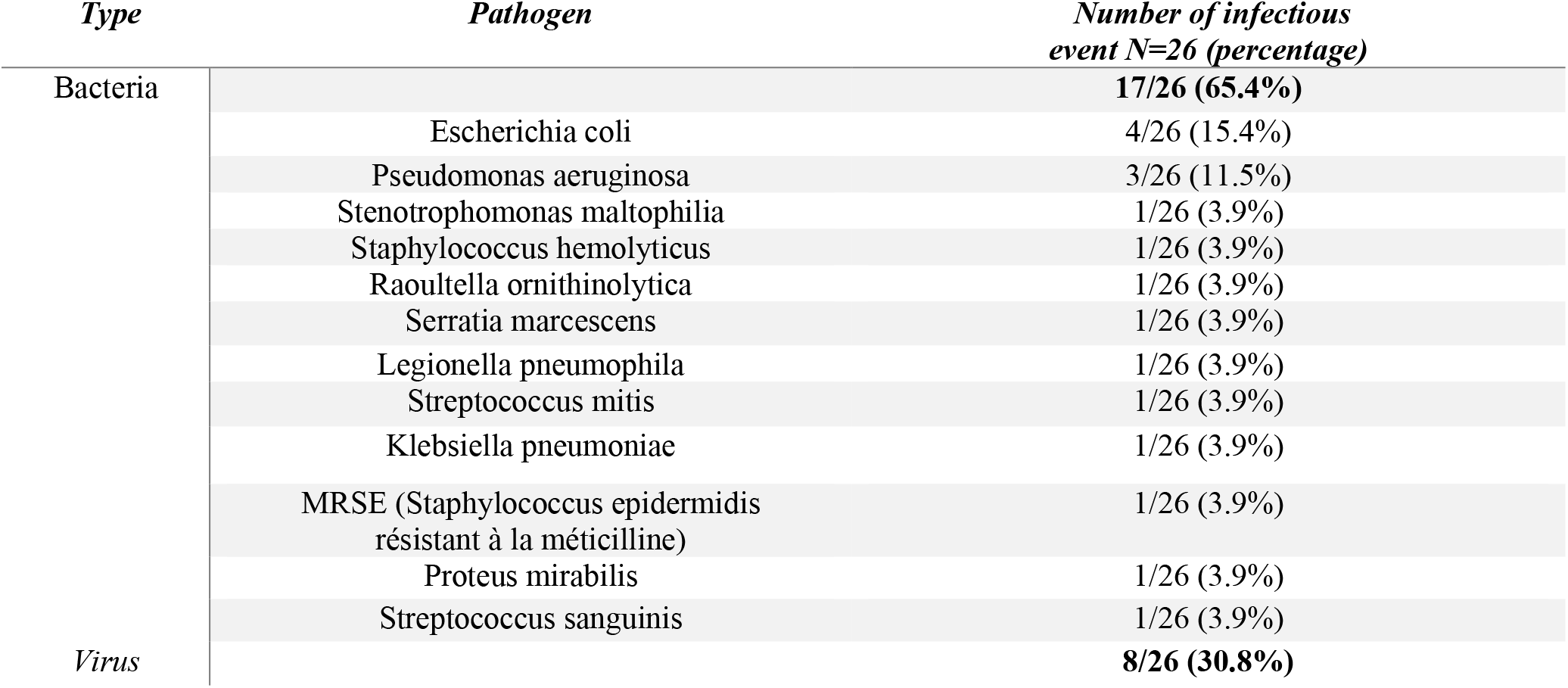
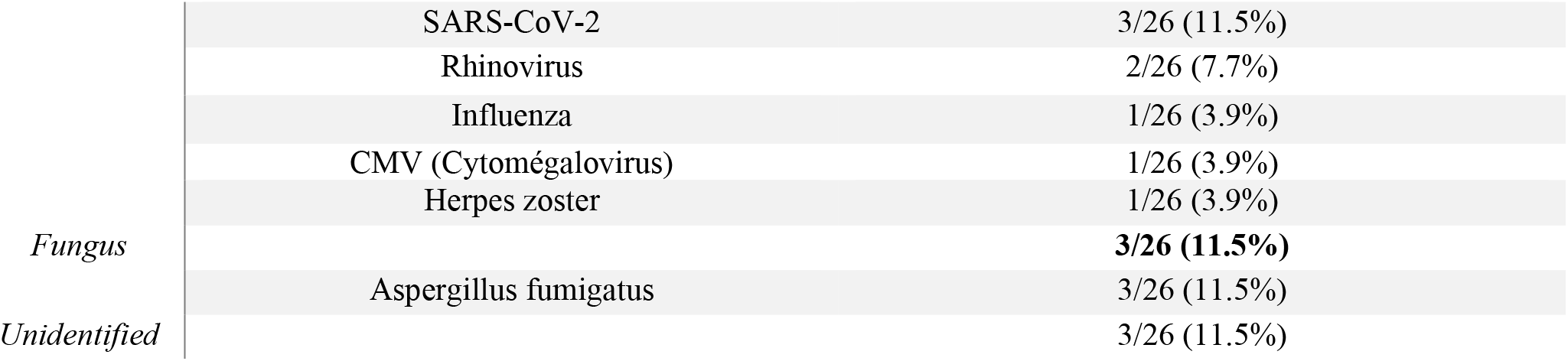
Proportions of the different pathogens in infectious events.

**Table 6.**
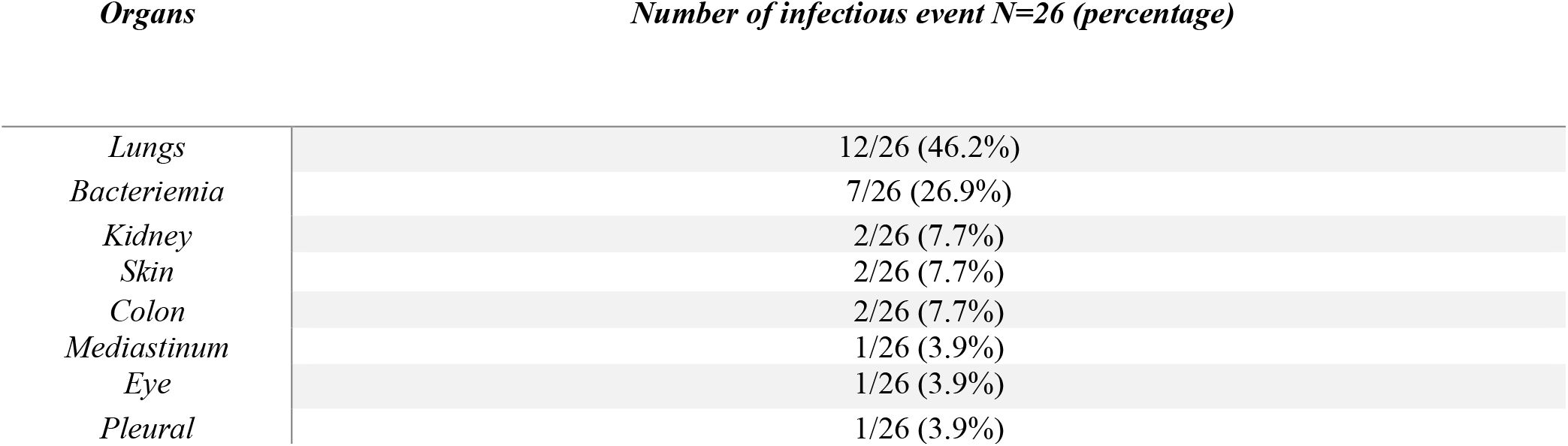
Proportion of different organ involvement in infectious events.

## Discussion

We present the first reported Belgian national cohort describing 21 cases of VEXAS syndrome across 4 third line Academic medical centers. All patients were male, consistent with previous reports (9). Our cohort shows clinical manifestations including general symptoms, cutaneous involvement, chondritis, articular, ophthalmic, and thromboembolic features, consistent with other cohorts (1,10). We report a lower frequency of pulmonary involvement than described in the literature. Nevertheless, it has previously been suggested that pulmonary involvement may be overestimated, particularly due to the high frequency of infections, which can sometimes be difficult to distinguish from a non-infectious inflammatory injury directedly linked to VEXAS, especially in the context of the SARS-CoV-2 pandemic concomitant with the initial description of the disease (7). Regarding cartilaginous involvement, only a few studies have reported costochondral involvement (11,12). Tracheal chondritis may be underestimated due to diagnostic difficulties, but it may account for the high prevalence of dyspnea and warrants further investigation (13). Concerning hematologic involvement, we report 11 cases of low-risk MDS without progression to high-risk MDS, which is Consistent to findings reported in the literature (14). The prevalence of MGUS remains low, in agreement with other cohorts (7). Bone marrow findings were also consistent with previously reported descriptions. Initial samples showed no blasts excess. Vacuolization and dysplasia were not systematically associated as well (15). Vacuoles were found in 70.6%of bone marrow samples (2,9). Incidence of vacuole findings compared to some other studies may be explained by the fact that some bone marrow samples were analyzed prior to 2020. At a time when the presence of vacuoles in myeloid progenitors was not systematically assessed, since VEXAS had not yet been described. Regarding cytopenias, a relatively low rate of macrocytic anemia at diagnosis was observed, consistent with the Swiss and Dutch cohorts (16,17). We observed one case of microcytic anemia due to concomitant thalassemia, which has not previously been reported outside our cohort (18). The low rate of genital, cardiac, renal, digestive, and neurological involvement is consistent with the literature (10). Concerning mutations, we described the canonical mutations affecting codon 41, as well as a rarer mutation involving codon 56.

Regarding treatment, administration of biologic therapies, in accordance with current consensus guidelines, were not constant in all patients. Given the limited access to biologic therapies in Belgium, they were prescribed in only 13 of our 17 living patients (76%). Three patients were treated with azacitidine. One of them did not present with myelodysplastic syndrome (MDS). This is already described in some other groups, as this drug can eliminate the mutant clone while simultaneously treating inflammation (19). We also describe two patients treated with allogeneic stem cell transplantation. Although this option may represent the best means of long-term disease control, it is associated with significant adverse effects. While it remains the only curative treatment, careful selection of candidates is essential. In our cohort, one transplanted patient developed multiple complications, including infections and graft-versus-host disease (GVHD), leading to death. Interestingly, one patient received only therapies targeting the mutant clone (lenalidomide and azacitidine), as he did not exhibit elevated C-reactive protein (CRP) levels and presented only with neutrophilic dermatosis and MDS. Five of our patients achieved “complete remission”: two under tocilizumab, one under azacitidine, one under ruxolitinib, and one following allogeneic stem cell transplantation. Fifty percent of patients treated with tocilizumab achieved complete remission, whereas only one patient (33%) treated with azacitidine did so, although all nearly reached remission without fully meeting our predefined criteria (5). These results highlight the effectiveness of these therapeutic approaches, as already documented in the literature (5). Nevertheless, in accordance with previously published data, all patients who received ruxolitinib or underwent transplantation achieved complete remission (16).

Regarding infections, we identified 26 distinct episodes affecting 12 patients. The spectrum of pathogens was less broad than that reported in the literature; notably, there was no Pneumocystis jirovecii or non-tuberculous mycobacterial infection (20). In contrast, we observed an overrepresentation of Legionella pneumophila, consistent with previous reports (7). This pathogen should therefore be systematically considered in cases of respiratory failure in patients with VEXAS syndrome. Among atypical pathogens, we observed pulmonary CMV infection, Raoultella ornithinolytica, and three cases of Aspergillus fumigatus infection. These opportunistic infections may highlight the immunodeficient component of VEXAS syndrome per se (7). Infectious prophylaxis remains a crucial issue, particularly in the context of opportunistic infections (20).

Our study has several limitations. First, the small sample size reduces statistical power and limits comparisons between groups. Second, as data collection forms were completed by multiple practitioners, variability in reporting and interpretation may have occurred. Finally, partial responses were not considered in this study, which may have limited the characterization of treatment responses. Last, we did not record all the belgian VEXAS cases diagnosed during the enrolling period.

## Conclusion

This study represents the first multicenter description of VEXAS syndrome in Belgium. Our results confirm the heterogeneous presentation of the syndrome and support the importance of screening for *UBA1* mutations in elderly men presenting with various autoinflammatory manifestations, particularly when associated with MDS. Second-line therapy in this cohort was heterogeneous, mainly involving anti–IL-6 receptor agents and JAK inhibitors, both recommended in the literature without a clearly defined hierarchy. Access to these treatments in Belgium is complicated by the lack of reimbursement for this indication. Nevertheless, these therapies were made available to patients in Belgium, predominantly through compassionate use programs or sample-based access mechanisms. We hope that our series may help raise awareness among reimbursement authorities regarding the unmet medical need for access to targeted therapies, which clinicians already advocate for in their practice, and which are supported by the most recent recommendations. At the present time, the main challenge remains infectious morbidity and mortality in patients with VEXAS syndrome. Consensus regarding optimal infectious prophylaxis strategies should be evaluated in larger studies.

## Data Availability

All data produced in the present work are contained in the manuscript

## Notes

### Competing Interest Statement

The authors have declared no competing interest.

### Author Declarations

This study was approved by the Ethics Committee of HUB (reference: P2025/015) as well as by the respective committees at each participating center.

